# Ability of a polygenic risk score to refine colorectal cancer risk in Lynch syndrome

**DOI:** 10.1101/2023.04.20.23288850

**Authors:** Nuria Dueñas, Hannah Klinkhammer, Nuria Bonifaci, Isabel Spier, Andreas Mayr, Emadeldin Hassanin, Anna Díez-Villanueva, Victor Moreno, Marta Pineda, Carlo Maj, Gabriel Capellá, Stefan Aretz, Joan Brunet

## Abstract

**Background:** Polygenic risk scores (PRS) have been used to stratify colorectal cancer (CRC) risk in the general population, whereas its role in Lynch syndrome (LS), the most common type of hereditary CRC, is still conflicting. We aimed to assess the ability of PRS to refine CRC risk prediction in European-descendant LS individuals.

**Methods:** 1,465 LS individuals (557 *MLH1*, 517 *MSH2/EPCAM*, 299 *MSH6*, and 92 *PMS2)* and 5,656 CRC-free population-based controls from two independent cohorts were included. A 91-Single Nucleotide Polymorphism PRS was applied. A Cox proportional hazard regression model with “family” as a random effect and a logistic regression analysis, followed by a meta-analysis combining both cohorts were conducted.

**Results:** Overall, we did not observe a statistically significant association between PRS and CRC risk in the entire cohort. Nevertheless, PRS was significantly associated with a slightly increased risk of CRC or advanced adenoma (AA), in those with CRC diagnosed < 50 years, and in individuals with multiple CRCs or AAs diagnosed < 60 years.

**Conclusion:** The PRS may slightly influence CRC risk in LS individuals, in particular in more extreme phenotypes such as early-onset disease. However, the study design and recruitment strategy strongly influence the results of PRS studies. A separate analysis by genes and its combination with other genetic and non-genetic risk factors will help refine its role as a risk modifier in LS.

**KEY MESSAGES:** WHAT IS ALREADY KNOWN ON THIS TOPIC?

- Great variability in the incidence of CRC has been described in LS individuals, even within the same family.
- Polygenic risk scores (PRS) can help stratify colorectal cancer risk and, thus, adjust surveillance or treatment procedures.

WHAT THIS STUDY ADDS

- PRS performed on family-based registries slightly influences CRC risk in subgroups of LS individuals, even though with weak effects.
- Our study showed a weak association of PRS with multiple and young CRC cases, pointing to a possible risk-modifying role in extreme phenotypes.

HOW THIS STUDY MIGHT AFFECT RESEARCH, PRACTICE OR POLICY

- Gene-based PRS analysis and its combination with other genetic and non-genetic factors may contribute to refining cancer risk in LS patients.

## INTRODUCTION

Colorectal cancer (CRC) is the third most incident cancer overall and the second leading cause of cancer-related death worldwide. Incidence rates are four times higher in the Global North, associated with lifestyle and dietary risk factors^1^.

About 5% of CRC is considered hereditary due to highly penetrant pathogenic germline variants in cancer-predisposing genes^2,3^. The main cause of hereditary CRC is Lynch Syndrome (LS), with an estimated carrier frequency in the general population of around 1:279^4^. It is characterised as an autosomal dominant inherited defect in any of the mismatch repair (MMR) genes (*MLH1, MSH2, MSH6, PMS2)* or *EPCAM* gene deletions, resulting in silencing of the *MSH2* gene in epithelial tissues^5^. Median CRC cumulative incidences at 75 years show an important variability according to mutated gene and gender: 48/57%, 47/51%, and 18/20% for male and female carriers of mutations in *MLH1, MSH2*, and *MSH6*, respectively, and 10% for both genders in carriers of mutations in *PMS2*^6^. Differences in CRC risk have also been identified based on the ethnic or geographical origin of carriers, with lower risks reported for European vs. American and Australasian individuals^7^. Moreover, LS carriers have an increased risk of developing multiple CRCs, CRC at a younger age, and other LS-associated cancers such as endometrial (EC) or ovarian cancer^6^.

In LS, as in other hereditary cancer predisposition syndromes characterised by incomplete penetrance, one of the main challenges is to identify which risk-modifying factors may modulate the expression of the cancer syndrome^7,8^. In recent years, multiple, common, low-penetrance CRC risk variants have been identified through genome-wide association studies (GWAS)^9–11^. Each risk allele individually confers a small risk, but their combined effect as a polygenic risk score (PRS) exhibits significant risks of developing CRC in the general population. Being in the highest PRS percentiles was shown to increase the risk of CRC two-to seven-fold^10,12–16^. Moreover, PRS might be particularly relevant in patients with a more extreme, i.e., severe, phenotype: a study performed in individuals diagnosed with CRC before 50 years of age (early-onset disease) demonstrated the existence of an interaction between PRS and CRC risk, with an odds ratio (OR) of 3.73 (3.28-4.24) in the highest PRS quartile^17^. Another study on familial CRC (individuals who fulfil Amsterdam or Bethesda criteria without a pathogenic germline MMR variant) identified an increased CRC risk in individuals in the highest 5% of the PRS distribution, with an OR of 4.89 (2.37-10.07)^18^.

To date, the modulating effect of PRS on CRC risk in LS individuals is still controversial. Two studies on a population-based repository from the UK Biobank (UKBB) including 76 and 388 LS carriers, respectively, reported that PRS may strongly influence CRC risk^16,19^; however, another analysis of the clinic-based registry of the Colon Cancer Family Registry (CCFR), including 826 European-descendant LS individuals, found no evidence of association, irrespective of sex or mutated gene^20^.

Our objective was to evaluate whether differences in CRC penetrance in European-descendant LS individuals can, in part, be explained by the accumulation of low-risk CRC alleles using a validated set of 91 SNPs for PRS analysis.

## METHODS

### Study participants

#### LS individuals

A total of 1,465 European-descendant individuals with genetically confirmed LS (557 *MLH1*, 517 *MSH2/EPCAM*, 299 *MSH6*, and 92 *PMS2)* from two independent cohorts were included: 918 LS individuals (353 families) identified at the Catalan Institute of Oncology (ICO; Spain) and 547 LS individuals (392 families) from the University Hospital of Bonn (UKB; Germany). Patients were recruited based on the fulfilment of Bethesda or Amsterdam criteria or via an EC and CRC-based LS screening programme (since 2016 at the ICO)^21^. Patients included were affected index patients and affected or unaffected carriers among the relatives identified through cascade testing. In the ICO LS cohort, there was a lower percentage of pathogenic *MSH2* variant carriers (mainly due to the existence of *MLH1* founder mutations in the ICO series) and a higher percentage of pathogenic *MSH6* and *PMS2* variant carriers (mainly identified through an EC/CRC-based LS screening) when compared with the UKB LS cohort. In addition, the ICO cohort included a higher proportion of non-index individuals. There were no significant differences in the distribution of affected genes between early-onset cases and the entire cohort ***(Table 1)***.

**TABLE 1.** Main characteristics of the population studied. *ICO*: Catalan Institute of Oncology, Spain; *UKB*: University Hospital of Bonn, Germany

|  | Lynch syndrome |  |  | Population controls |  |  |
| --- | --- | --- | --- | --- | --- | --- |
|  | All series | ICO | UKB | All series | Spain | Germany |
| **Total n of individuals** | **1465**<br>**(100%)** | 918<br>(62.7%) | 547<br>(37.3%) | **5656**<br>**(100%)** | 1642<br>(29.0%) | 4014<br>(70.1%) |
| **Age** |  |  |  |  |  |  |
| Mean age at censor (range) | **45.6 (12-93)** | 47.9 (16-93) | 41.6 (12-86) | **71.0 (24-94)** | 62.4 (24-92) | 74.5 (49-94) |
| **Gender** |  |  |  |  |  |  |
| Male | **707**<br>**(48.3%)** | 409<br>(44.6%) | 298<br>(54.5%) | **2813**<br>**(49.7%)** | 835<br>(50.9%) | 1978<br>(49.3%) |
| Female | **758**<br>**(51.7%)** | 509<br>(55.4%) | 249<br>(45.5%) | **2843**<br>**(50.3%)** | 807<br>(49.2%) | 2036<br>(50.7%) |
| **Mutated gene** |  |  |  |  |  |  |
| *MLH1* | **557**<br>**(38.0%)** | 346<br>(37.7%) | 211<br>(38.6%) | - | - | - |
| *MSH2/EPCAM* | **517**<br>**(35.3%)** | 247<br>(26.9%) | 270<br>(49.4%) | - | - | - |
| *MSH6* | **299**<br>**(20.4%)** | 250<br>(27.2%) | 49 (9.0%) | - | - | - |
| *PMS2* | **92**<br>**(6.3%)** | 75<br>(8.2%) | 17<br>(3.1%) | - | - | - |
| **Index case** |  |  |  |  |  |  |
| Yes | **590**<br>**(40.3%)** | 193<br>(21.0%) | 397<br>(72.6%) | - | - | - |
| No | **875**<br>**(59.7%)** | 725<br>(79.0%) | 150<br>(27.4%) | - | - | - |

All patients gave informed consent and the internal Ethics Committee approved this study.

#### Non-LS individuals

A total of 5,656 unselected CRC-free individuals from the same population were included in the analysis (CRC-free population controls): 1,642 individuals from Spain and 4,014 from Germany. The controls from Spain included individuals from the CRCGEN study and individuals participating in a population-based CRC screening program, most of whom had a positive faecal immunochemical test (FIT) result and a colonoscopy without cancer or advanced adenoma, as described elsewhere^22^. The German controls were drawn from the population-based Heinz Nixdorf RECALL (HNR) study (Risk Factors, Evaluation of Coronary Calcification, and Lifestyle) as described recently^23^ ***(Table 1)***.

### Data collection

Clinical data included demographic, personal and oncologic history, and follow-up carried out from birth to 06/2021. In LS individuals, histories of colorectal polyps or other LS-related cancers were also collected. Data supporting the results were stored in local databases at both centres.

### SNP selection

The selected SNPs (n=95) and associated risks were obtained from the meta-analysis of CRC risk alleles performed by Huyghe *et al*.,^10^ ***(Tab. S1)*** and were commonly used to study sporadic CRC risk at the initiation of the study^16,19^. Individual CRC risk-associated SNPs reached independent genome-wide significance (p<5×10^−8^) in a large-scale GWAS.

### Genotyping

ICO blood DNA samples were genotyped with the Illumina Global Screening Array-24 (GSA) v2.0 and v3.0 (https://emea.illumina.com/science/consortia/human-consortia/global-screening-consortium.html) and UKB samples with GSA v3.0. Of note, 48% of the ICO population of CRC-free individuals were previously included in the meta-analysis by Huyghe *et al*.,^10^ however, they corresponded to ∽1% of the total number of cases and controls in the analysis. Details regarding quality control procedures and correlation between arrays have been described previously^18,22^.

Non-European-descendant individuals were excluded from the analysis. To assess ethnicity, Spanish samples were compared with 1,397 HapMap samples, while German samples were compared with 1k genome samples. Classification into different ethnicity groups was performed by selecting ancestry-informative marker SNPs (AIM SNPs) and using a principal components analysis (PCA) approach.

### Imputation

Thirteen and eighteen of the 95 variants of interest were included in the Illumina GSA-24 v2.0 and v3.0, respectively. Variants not directly genotyped by the corresponding arrays were imputed in the ICO with the Michigan Imputation Server (HRC version r1.1.2016 panel)^24^ and in the UKB with a comparable pipeline based on the bioinformatic tools bcftools, minimac, and vcftools, using GRCh37 as the reference genome (1000 Genomes, phase 3, v5)^25^. Missing variants and variants with an imputation quality (r^2^) <0.3 (considering all genotyped samples) were not included in the final PRS analysis, which resulted in the exclusion of rs6058093, rs35470271, rs145364999, and rs755229494 ***(Tab. S1)***.

### PRS calculation

For each participant, PRS was computed using the PLINK score function^26^, based on the 91 quality-controlled CRC risk alleles (coded as 0, 1, or 2) and effect sizes as reported by Huyghe *et al*., (PRS) and averaged over the number of observed variants per individual^10^ (wPRS). To ease interpretation, wPRS values were rescaled (rwPRS) to indicate risk per allele (using the ratio of non-averaged PRS and wPRS values in controls as a scaling factor) as previously reported^18^.

### Study events

Two events were considered: i) CRC and ii) advanced adenoma (AA) (adenoma with significant villous features (>25%), size ≥1.0 cm, high-grade dysplasia, or early invasive cancer).

Two subgroups were defined for the primary analysis: affected individuals (CRC and CRC or AA LS individuals) and unaffected individuals (CRC-free or CRC-free and AA-free LS individuals). For the subanalysis of multiple CRCs, three subgroups were defined: multiple events (multiple CRC and multiple CRC or AA LS individuals), single event (single CRC and single CRC or AA LS individuals) and no-event (CRC-free and CRC-free and AA-free LS individuals). CRC-free population controls were only compared to CRC or multiple CRC LS individuals when considering CRC as a study event as no reliable information was available regarding AA in this population. ***(Tab. S2, Tab. S3, and Fig. S1)***.

### Statistical methods

Statistical analyses and graphical representations were conducted with R version 4.0.5. For the primary analysis, the association of rwPRS with CRC and CRC or AA risk was tested by considering time to CRC (years since birth to event of study) using a Cox proportional hazard regression model with family as a random effect (frailty model). Observations in the control cohort were right censored at the age of last contact and CRC diagnosis (yes/no) was used as an event variable. The date of the first polypectomy for adenoma was used as a time-dependent variable. Additionally, sex, birth cohort (<1940, 1940-49, 1950-59, 1960-69, 1970-79, >1980), and other LS-related cancers were included as covariates.

For the subanalysis of multiple CRCs, the association of rwPRS with multiple CRCs or AAs was tested using a mixed effects logistic regression, including age, sex, birth cohort, polypectomy before the second CRC, the occurrence of other cancers, and family (random effect) as covariates.

Results from both cohorts (ICO and UKB) were combined and analysed via a fixed-effect meta-analysis and the inverse-variance method. The combined rwPRS effect was estimated as the weighted average of the estimates of the individual studies and weights were derived as the inverse of the variance of the individual effect estimate. The population was stratified according to rwPRS tertiles using the medium category as a reference. Additionally, to test for heterogeneity, Cochran’s Q was computed on the derived estimates and a χ^2^-test with one degree of freedom was performed. Results with *p*-values <0.05 in the test for heterogeneity were not considered. The meta-analysis was conducted via R package meta^27^. To correct for multiple testing, analyses were grouped by study event and control group, and *p*-values inside these groups were corrected via false discovery rate (FDR) correction^28^. Only results with *p*-values <0.05 after FDR correction (*p*-FDR) were considered statistically significant.

## RESULTS

No differences in PRS distribution were observed when comparing CRC-free LS individuals and CRC-free population controls in any of the cohorts studied ***(Fig. S2)***.

### Primary analysis

#### CRC as the study event

A statistically significant association of rwPRS with CRC risk was found in LS carriers under 50 years of age compared with CRC-free LS individuals (HR=1.022 [1.007-1.038], *p*-FDR=0.01). We found a tendency for an association of rwPRS with CRC risk in the entire cohort and *MSH6* variant carriers. We found no statistically significant association of rwPRS with CRC risk when comparing CRC LS to CRC-free population individuals ***(Table 2 and Tab. S4)***.

**TABLE 2.**
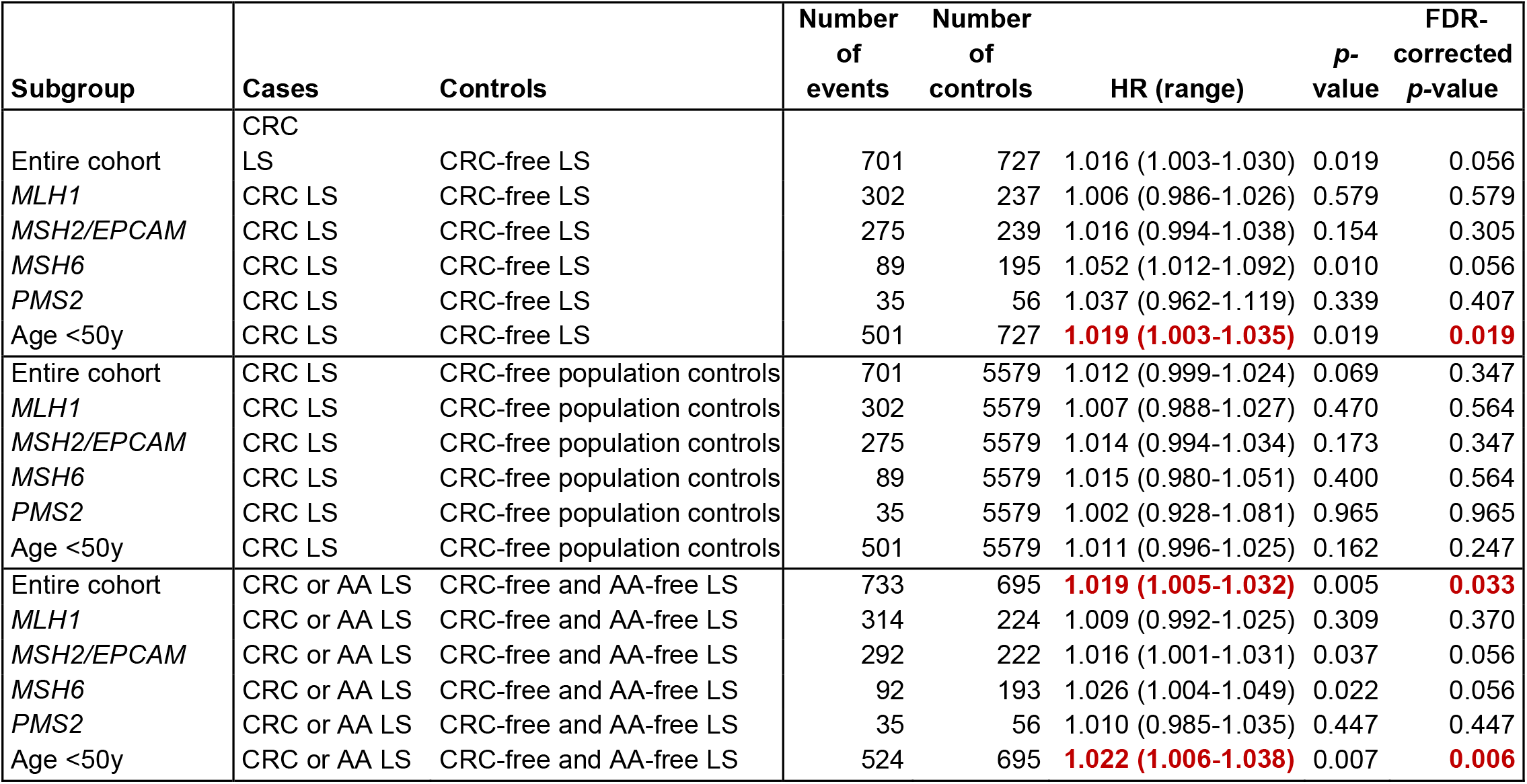
Meta-analysis results of the association between Polygenic risk score (rwPRS) and colorectal cancer (CRC) risk or CRC and advanced adenoma (AA) risk. *CRC* LS: LS individuals previously diagnosed of CRC; *CRC-free* LS: LS individuals with no previous CRC diagnosis; CRC-free population control: individuals from the population without previous diagnosis of CRC; *CRC or AA* LS: LS individuals with a previous diagnosis of CRC or AA, whichever occurred first; *CRC-free and AA-free* LS: LS individuals not diagnosed with CRC or AA; *HR*: Hazard ratio; *FDR* False discovery rate; *age <50y*: cases with CRC <50 years of age.

Additionally, rwPRS tended to be associated with higher CRC risk in *MSH2/EPCAM* (tertile low: HR=0.716 [0.505-1.016], *p*-FDR= 0.53 vs. tertile high: HR=1.058 [0.769-1.455] *p*-FDR=0.96) and *MSH6* variant carriers (tertile low: HR=0.617 [0.299-1.271], *p*-FDR=0.53 vs. tertile high: HR=1.594 [0.929-2.735], *p*-FDR=0.53), however, results were not statistically significant ***(Figure 1)***.

**FIGURE 1.**
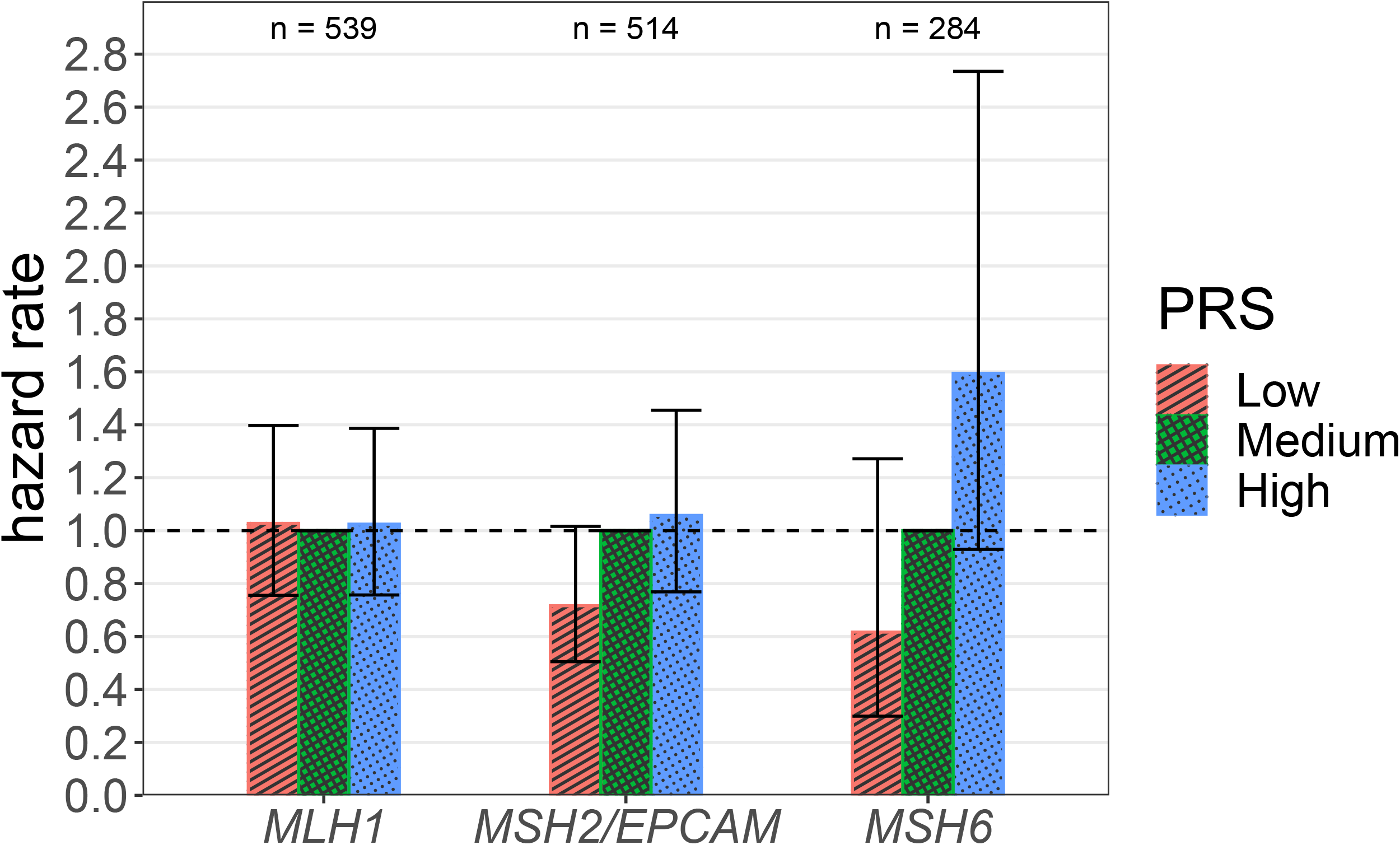
Interplay of each mutated germline gene and polygenic risk score (rwPRS) for colorectal cancer (CRC) risk. CRC risk for each mutated gene, stratified according to rwPRS tertiles using the intermediate rwPRS tertile as the reference group. 95% confidence intervals are indicated by vertical lines. PMS2 carriers are not included due to the low sample size.

#### CRC or AA as study events

A statistically significant association of rwPRS with CRC or AA risk was observed in the entire cohort (HR=1.019 [1.005-1.032], *p-*FDR=0.03) and in LS carriers under 50 years of age (HR=1.022 [1.006-1.038], *p-*FDR=0.006). We observed a tendency for an association of rwPRS with CRC or AA risk in *MSH2/EPCAM* and *MSH6* carriers ***(Table 2 and Tab. S5)***.

Even though no statistically significant associations were observed ***(Figure 2)***, rwPRS tended to be associated with a higher risk of CRC and AA in *MSH6* variant carriers (tertile low: HR=0.669 [0.322-1.393], *p-*FDR=0.57 vs. tertile high: HR=2.015 [1.169-3.471], *p-*FDR=0.39).

**FIGURE 2.**
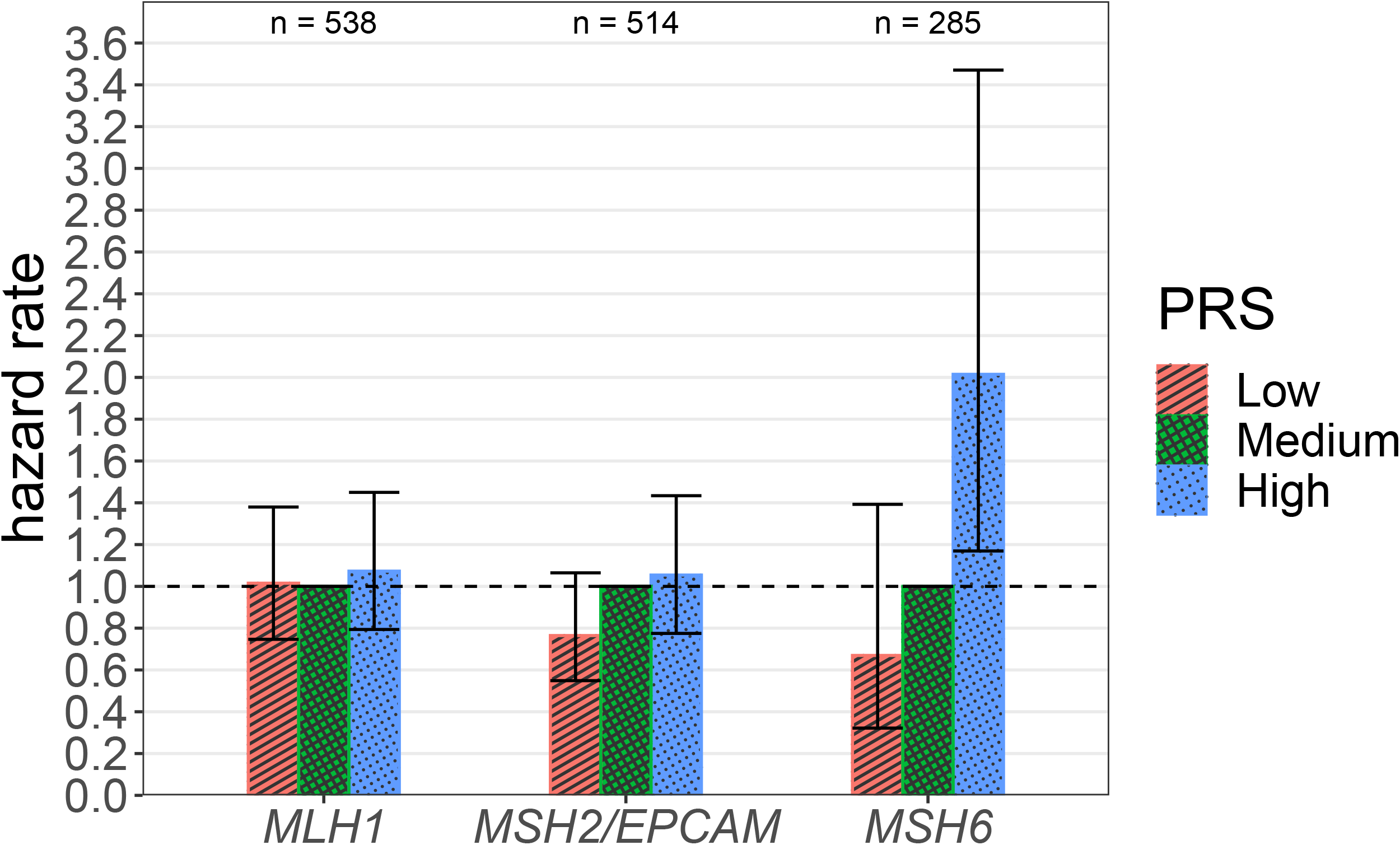
Interplay of each mutated germline gene and polygenic risk score (rwPRS) for colorectal cancer (CRC) or advanced adenoma (AA) risk. Risk of CRC or AA for each mutated gene, stratified according to rwPRS tertiles using the intermediate rwPRS tertile as the reference group. 95% confidence intervals are indicated by vertical lines. PMS2 carriers are not included due to the low sample size.

### Subanalysis: multiple CRCs

#### Multiple CRCs as the study event

No statistically significant association of rwPRS with multiple CRC risk was observed (irrespective of the gene involved) when comparing single-CRC LS cases, CRC-free LS individuals, or CRC-free population controls ***(Table 3 and Tab. S6)***. These analyses could not be performed in *MSH6* or *PMS2* carriers due to the low sample sizes.

**TABLE 3.**
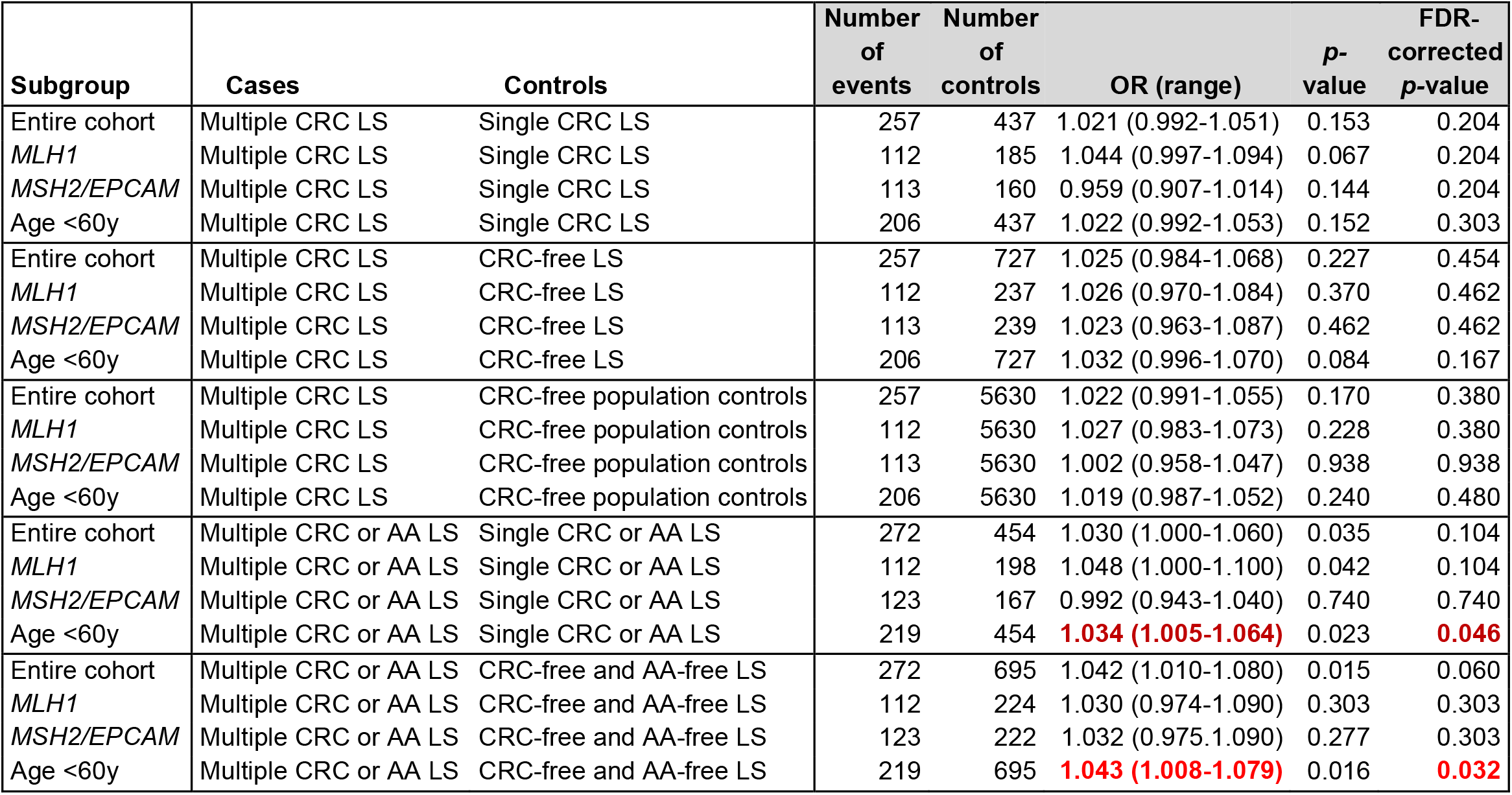
Meta-analysis results of the association between polygenic risk score (rwPRS) and multiple colorectal cancer (CRC) risk or multiple CRC or advanced adenoma (AA) risk. s*Multiple CRC LS*: LS individuals previously diagnosed with more than one CRC; *Single CRC LS*: LS individuals previously diagnosed with one CRC; *CRC-free LS*: LS individuals without previous diagnosis of CRC; *CRC-free population*: individuals from the population without previous diagnosis of CRC; *Multiple CRC or AA LS*: LS individuals previously diagnosed of multiple CRCs, multiple AAs or at least one CRC and one AA; *Single CRC or AA LS*: LS individuals previously diagnosed of CRC or AA, whichever occurred first; *CRC-free and AA-free LS*: LS individuals without previous diagnosis of CRC or AA; *OR*: Odds ratio; *FDR*: False discovery rate; *Age <60y*: cases with CRC <60 years of age.

| Subgroup | Cases | Controls | Number of events | Number of controls | OR (range) | p-value | FDR-corrected p-value |
| --- | --- | --- | --- | --- | --- | --- | --- |
| Entire cohort | Multiple CRC LS | Single CRC LS | 257 | 437 | 1.021 (0.992-1.051) | 0.153 | 0.204 |
| *MLH1* | Multiple CRC LS | Single CRC LS | 112 | 185 | 1.044 (0.997-1.094) | 0.067 | 0.204 |
| *MSH2/EPCAM* | Multiple CRC LS | Single CRC LS | 113 | 160 | 0.959 (0.907-1.014) | 0.144 | 0.204 |
| Age <60y | Multiple CRC LS | Single CRC LS | 206 | 437 | 1.022 (0.992-1.053) | 0.152 | 0.303 |
| Entire cohort | Multiple CRC LS | CRC-free LS | 257 | 727 | 1.025 (0.984-1.068) | 0.227 | 0.454 |
| *MLH1* | Multiple CRC LS | CRC-free LS | 112 | 237 | 1.026 (0.970-1.084) | 0.370 | 0.462 |
| *MSH2/EPCAM* | Multiple CRC LS | CRC-free LS | 113 | 239 | 1.023 (0.963-1.087) | 0.462 | 0.462 |
| Age <60y | Multiple CRC LS | CRC-free LS | 206 | 727 | 1.032 (0.996-1.070) | 0.084 | 0.167 |
| Entire cohort | Multiple CRC LS | CRC-free population controls | 257 | 5630 | 1.022 (0.991-1.055) | 0.170 | 0.380 |
| *MLH1* | Multiple CRC LS | CRC-free population controls | 112 | 5630 | 1.027 (0.983-1.073) | 0.228 | 0.380 |
| *MSH2/EPCAM* | Multiple CRC LS | CRC-free population controls | 113 | 5630 | 1.002 (0.958-1.047) | 0.938 | 0.938 |
| Age <60y | Multiple CRC LS | CRC-free population controls | 206 | 5630 | 1.019 (0.987-1.052) | 0.240 | 0.480 |
| Entire cohort | Multiple CRC or AA LS | Single CRC or AA LS | 272 | 454 | 1.030 (1.000-1.060) | 0.035 | 0.104 |
| *MLH1* | Multiple CRC or AA LS | Single CRC or AA LS | 112 | 198 | 1.048 (1.000-1.100) | 0.042 | 0.104 |
| *MSH2/EPCAM* | Multiple CRC or AA LS | Single CRC or AA LS | 123 | 167 | 0.992 (0.943-1.040) | 0.740 | 0.740 |
| Age <60y | Multiple CRC or AA LS | Single CRC or AA LS | 219 | 454 | **1.034 (1.005-1.064)** | 0.023 | **0.046** |
| Entire cohort | Multiple CRC or AA LS | CRC-free and AA-free LS | 272 | 695 | 1.042 (1.010-1.080) | 0.015 | 0.060 |
| *MLH1* | Multiple CRC or AA LS | CRC-free and AA-free LS | 112 | 224 | 1.030 (0.974-1.090) | 0.303 | 0.303 |
| *MSH2/EPCAM* | Multiple CRC or AA LS | CRC-free and AA-free LS | 123 | 222 | 1.032 (0.975-1.090) | 0.277 | 0.303 |
| Age <60y | Multiple CRC or AA LS | CRC-free and AA-free LS | 219 | 695 | **1.043 (1.008-1.079)** | 0.016 | **0.032** |

#### Multiple CRCs or AAs as study events

A significant association of rwPRS with multiple CRC or AA risk was observed in LS individuals under 60 years when comparing with single-CRC or AA LS (HR=1.057 [1.010-1.100], *p-*FDR=0.04) and CRC-free and AA-free LS (HR=1.043 [1.008-1.079], *p-* FDR=0.03). A tendency was observed for an association of rwPRS with multiple CRC or AA risk in the entire cohort and *MLH1* carriers ***(Table 3 and Tab. S7)***. These analyses could not be performed in *MSH6* and *PMS2* carriers due to the low sample size.

## DISCUSSION

PRS is regarded as an important addition to the assessment of an individual’s genetic risk in patients with sporadic and hereditary cancers; it can be used to identify individuals with a CRC risk several times lower or higher than that of the average population. In this way, its implementation seems to be a promising approach for a more individualised risk stratification. Several studies described the impact of PRS on modelling CRC risk in the general population^10,12–16^. In line with this, the risk alleles of those SNPs were found to accumulate in unexplained familial and early-onset CRC cases^17,18^. However, the interplay between a PRS based on sporadic CRC-associated SNPs and LS CRC risk remains controversial.

It is well known that among patients with hereditary CRC, in particular Lynch syndrome, the age of onset and cumulative CRC incidence is very heterogeneous, even within the same family transmitting the same pathogenic germline variant^6^. The estimated gene-specific, individual lifetime CRC risks of LS patients with *MLH1* or *MSH2* variants can be lower than 10% or as high as 90%-100% in a considerable fraction, highlighting relevant genetic and non-genetic modifiers of CRC risk^7,8^. Initially, a small subset of common CRC-associated SNPs was analysed in selected LS cohorts^29–32^. More recently, some studies used a more comprehensive set of around 100 CRC-associated SNPs in large population-based or familial CRC cohorts with conflicting results^16,19,20^.

Herein, we used a large, combined cohort of 1,465 affected and unaffected LS patients with pathogenic MMR germline variants, recruited at two European centres based on the fulfilment of clinical criteria (revised Bethesda or Amsterdam criteria) or as a result of an EC or CRC-based LS screening programme, to evaluate to what extent the polygenic background modulates CRC risk. When we compared LS carriers with CRC against population-based CRC-free controls (mean age 71 years), we did not observe any significant effect of PRS on CRC risk, neither in the entire cohort nor in subgroups (gene-specific groups, early-onset group). Nevertheless, the PRS was associated with a modestly increased risk of CRC or AA in the entire LS cohort. These results are in line with the work by Jenkins *et al*., which is based on a similar study design, recruitment strategy, and a set of 107 SNPs used for PRS calculation^20^. In that work, 826 European-descent LS carriers from the Colon Cancer Family Registry (CCFR) were included and the authors found no statistical evidence of an association between PRS and CRC risk, irrespective of sex or mutated gene.

Regarding the analysis between CRC and CRC-free LS probands, we did not find a statistically significant association between CRC and PRS in the entire cohort or the different subgroups except for early-onset LS CRC cases (<50 years) and LS with multiple CRCs or AAs (< 60 years), where a slightly increased CRC risk was evidenced. In contrast, two recent studies using UKBB data and the same 95 SNPs for PRS calculation, demonstrated that the polygenic background substantially influences CRC risk in LS with ORs ranging from 8 to 118 (estimated effect of PRS), or 4 to 16 (calculated effect of PRS) compared with the median tertile of the CRC-free population^16,19^. According to these results, PRS would account for parts of the interindividual variation in CRC risk among LS carriers and might contribute towards a clinically relevant individualised risk stratification.

The most obvious explanation for the apparently discrepant results between family-based (Jenkins *et al*.,^20^ and the present study) and population-based (Fahed *et al*.,^16^ and Hassanin *et al*.,^19^) studies is differences in study design and recruitment strategies. The LS probands from the two familial CRC registry studies were mainly recruited based on established clinical criteria, in particular early-onset and familial clustering of CRC and other LS-related tumours. Consistent with this ascertainment approach, the vast majority of participants carry pathogenic variants in the highly penetrant *MLH1* and *MSH2* genes **(Table 4)**, which are likely to be less influenced by the genetic background.

**TABLE 4.**
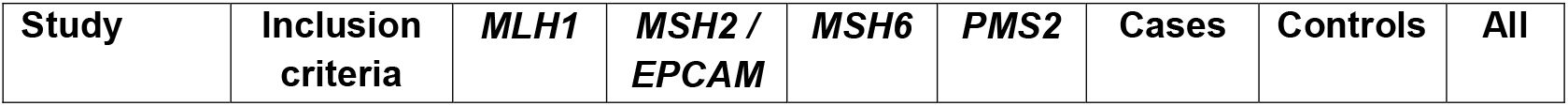
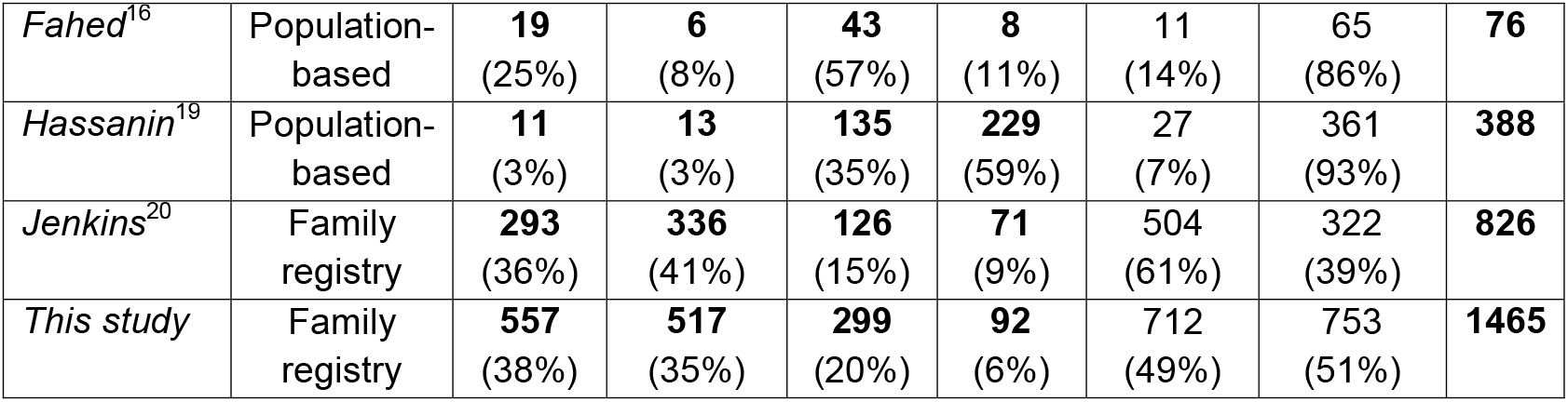
Characteristics of inclusion criteria and distribution of affected MMR genes in the different studies analysing PRS in LS individuals.

In contrast, studies using individuals from a population-based repository (UKBB) show a different distribution of affected MMR genes, with the vast majority of LS individuals carrying pathogenic variants in the moderate and low penetrance genes *MSH6* or *PMS2* **(Table 4)**. In a gene-specific analysis, Hassanin *et al*., found that the modifying effect of the PRS is inversely related to the penetrance of the MMR gene, with the strongest effect in *MSH6* and *PMS2* carriers^19^, which are clearly underrepresented in the studies of Jenkins *et al*.,^20^ and the present.

This is in line with hereditary breast cancer, where PRS has proven most relevant as a cancer risk modifier in carriers of pathogenic variants in moderate penetrance genes such as *CHEK2, ATM*, or *PALB2* compared with *BRCA1/2*^33,34^. While it can be expected that PRS may have a major influence in less penetrant CRC risk genes, we have not been able to show a significant effect, likely due to the small numbers of *MSH6* and *PMS2* variant carriers present in our family-based cohorts due to the aforementioned selection bias.

Another plausible explanation for the differences observed may be the sample size. In this study, we included twice as many LS individuals as Jenkins *et al*.^20^ and four and 20 times more than the UKBB analyses^16,19^. Moreover, the distribution and composition of cases and controls differ between family-registry and population-based studies **(Table 4)**. In Jenkins *et al*., and our study, the percentages of affected (LS carriers with CRC) and unaffected (CRC-free LS carriers) individuals are similar and some are members of the same family. Hence, the CRC-free LS controls are relatives of the cases and, thus, they likely share parts of the polygenic background and other risk factors with their affected relatives (cases) to a certain extent, which may explain the observed missing effect of PRS. In contrast, the UKBB studies include ten times more controls, supposedly healthy LS carriers apparently unrelated to the CRC LS cases. In this regard, it was shown that both in sporadic CRC^35^ and LS CRC^19^, family history and PRS are largely independent and provide complementary information about CRC risk.

On the other hand, there are differences in the results obtained between Jenkins *et al*.,^20^ and the present study. These differences can be explained by the sample size, as discussed above, and methodological differences. LS individuals in Jenkins *et al*., were censored after a polypectomy, while we considered the first polypectomy as a time-dependent variable of CRC risk, as per studies showing a reduction in CRC incidence in LS individuals undergoing regular colonoscopies^36,37^. However, since alternative pathways of colorectal carcinogenesis seem to exist in LS carriers, which originate doubts regarding the risk-reducing impact of colonoscopies, especially in *MLH1* carriers, future evidence will determine whether it is useful to apply this time-dependent variable correction in LS individuals^38,39^. Taken together, this and previous PRS studies on LS demonstrate that the study design and recruitment strategy strongly influence the results and conclusions of the PRS.

Finally, when we analysed extreme phenotypes, such as early-onset CRC (<50y) and young (<60y) LS cases with multiple CRCs or AAs, a significant, albeit low, association between PRS and risk was observed, pointing to a possible contribution of PRS to these higher-risk situations.

Some authors questioned whether the same CRC-associated SNPs identified in the general population and their specific effect sizes can be applied to stratify CRC risk in LS individuals and whether both specific SNPs and their risk-modifying power may differ for each mutated gene^8,20,29^. The potential identification of LS risk-modifier SNPs in large GWASs might contribute to the description of more specific risk-modulating factors in the future.

PRS studies in much larger, international, multicentric, LS cohorts are needed to more precisely estimate the PRS effect size in LS individuals, especially in those with extreme phenotypes, to evaluate the relevance of the polygenic background and interplay with other genetic and non-genetic risk factors. This will enable its eventual application in routine clinical practice.

In summary, this work shows, for the first time in a family-registry LS cohort, that the PRS can influence the CRC risk in specific subgroups of LS individuals, albeit with very weak effect sizes, which contrasts with the clearer modulating effect of the PRS in LS carriers identified in population-based cohorts.

## Data Availability

All data produced in the present study are available upon reasonable request to the authors

## ACKNOWLEDGEMENTS

We thank the participating patients and families and all members of the Units of Genetic Counseling and Genetic Diagnostic of the Hereditary Cancer Program of the Catalan Institute of Oncology (ICO-IDIBELL) and the Institute of Human Genetics of the University Hospital Bonn as well as the BufaLynch association for their support and funding of ICO’s Lynch Syndrome Database. We thank Gemma Aiza for technical support. The authors would also like to acknowledge the Department of Medicine at the Universitat Autònoma de Barcelona and the CERCA Program/Generalitat de Catalunya for institutional support.

## COMPETING INTERESTS

The authors declare no conflicts of interest.

## FUNDING

This research has been funded by the Instituto de Salud Carlos III and co-funded by the European Social Fund—ESF investing in your future (grant CM19/00099), the Catalan-Balearic Society of Oncology (2018 grant of the Catalan-Balearic Society of Oncology), the European Union’s Horizon 2020 research and innovation program under the EJP RD COFUND-EJP nº 825575, the Spanish Ministry of Economy and Competitiveness and the Spanish Ministry of Science and Innovation, co-funded by FEDER funds -a way to build Europe-(grants SAF2015-68016-R and PID2019-111254RB-I00), CIBERONC (CB16/12/00234) and the Government of Catalonia (SGR_01112). ADV and VM are part of group 55 of CIBERESP. The GSA genotyping was performed at the Spanish National Cancer Research Centre, in the Human Genotyping lab, a member of CeGen, PRB3, and is supported by grant PT17/0019, of the PE I+D+i 2013-2016, funded by ISCIII and ERDF. ADV was supported by PERIS contract SLT017/20/000042. This research was partially funded by public grants from the Spanish Association Against Cancer (AECC) Scientific Foundation—grant GCTRA18022MORE—and Spanish Ministry for Economy and Competitivity, Instituto de Salud Carlos III, co-funded by FEDER funds –a way to build Europe (FIS PI14-00613).

## AUTHOR CONTRIBUTIONS

<u>Núria Dueñas</u> – Study concept and design, Analysis and interpretation of data, Drafting of the manuscript, Critical revision of the manuscript for important intellectual content

<u>Hannah Klinkhammer, Nuria Bonifaci, Anna Diez-Villanueva</u> – Study concept and design, Statistical analysis, Analysis and interpretation of data, Drafting of the manuscript, Critical revision of the manuscript for important intellectual content

<u>Andreas Mayr, Emadeldin Hassanin, Carlo Maj</u> – Study concept and design, Statistical analysis, Analysis and interpretation of data, Critical revision of the manuscript for important intellectual content

<u>Anna Díez-Villanueva, Victor Moreno</u> – Study concept and design, Critical revision of the manuscript for important intellectual content

<u>Isabel Spier, Marta Pineda, Gabriel Capellá, Stefan Aretz, Joan Brunet</u> – Study concept and design, Analysis and interpretation of data, Critical revision of the manuscript for important intellectual content, Study supervision

## Notes

### Competing Interest Statement

The authors have declared no competing interest.

### Author Declarations

Ethics Committee of the Bellvitge University Hospital gave ethical approval for this work

